# Early versus late third trimester maternal SARS-CoV-2 BNT162b2 mRNA immunization maximizes transplacental antibody transfer and neonatal neutralizing antibody levels

**DOI:** 10.1101/2021.08.30.21262875

**Authors:** Amihai Rottenstreich, Gila Zarbiv, Esther Oiknine-Djian, Olesya Vorontsov, Roy Zigron, Geffen Kleinstern, Dana G. Wolf, Shay Porat

**Author notes:** **Funding:** No external funding was used for this study. **Corresponding Authors:** Amihai Rottenstreich, MD, Department of Obstetrics and Gynecology, Hadassah-Hebrew University Medical Center, POB 12000, Jerusalem, Israel, 91120., Dana G. Wolf, MD, Clinical Virology Unit, Department of Clinical Microbiology and Infectious Diseases, Hadassah-Hebrew University Medical Center, Jerusalem, Israel, 91120.

## Abstract

**Objective:** We aimed to assess the impact of early versus late third trimester maternal SARS-CoV-2 vaccination on transplacental transfer and neonatal levels of SARS-CoV-2 antibodies.

**Methods:** Maternal and cord blood sera were collected following term delivery after antenatal SARS-CoV-2 BNT162b2 mRNA vaccination, with the first vaccine dose administered during 27-36 weeks gestation. SARS-CoV-2 spike protein (S) and receptor binding domain (RBD)- specific, IgG levels and neutralizing potency were evaluated in maternal and cord blood samples.

**Results:** The study cohort consisted of 171 parturients (median age, 31 years; median gestational age, 39.7 weeks): 83 (48.5%) immunized at early 3^rd^ trimester (1^st^ dose at 27-31 weeks), and 88 (51.5%) immunized at late 3^rd^ trimester (1^st^ dose at 32-36 weeks). All mother-infant paired sera were positive for anti S- and anti-RBD-specific IgG. Anti-RBD-specific IgG concentrations in neonatal sera were higher following early versus late 3^rd^ trimester vaccination and were positively correlated with increasing time since vaccination (r=□0.26; P=0.001). The median placental transfer ratios of anti-S and anti-RBD specific IgG were increased following early versus late 3^rd^ trimester immunization (anti-S ratio:1.3 vs. 0.9, anti-RBD-specific ratio:2.3 vs. 0.7, P<0.001). Neutralizing antibodies placental transfer ratio was greater following early versus late 3^rd^ trimester immunization (1.9 vs. 0.8, P<0.001), and was positively associated with longer duration from vaccination (r=□0.77; P<0.001).

**Conclusions:** Early- as compared to late third trimester maternal SARS-CoV-2 immunization enhanced transplacental antibody transfer and increased neonatal neutralizing antibody levels. Our findings highlight that vaccination of pregnant women early in the third trimester may optimize neonatal seroprotection.

## Introduction

On March 2020, Coronavirus Disease 19 (COVID-19) caused by severe acute respiratory syndrome coronavirus 2 (SARS-CoV-2), was declared a pandemic by the World Health Organization [1]. Up to May 2021, the pandemic has afflicted over 152 million individuals resulting in over 3 million deaths. Global efforts to combat COVID-19 have led to the development of numerous vaccines including two novel mRNA-based vaccines, which were shown to be highly effective in preventing SARS-CoV-2-related illness [2-4]. In Israel, a nationwide mass vaccination campaign against COVID-19 using the BNT162b2 (Pfizer/BioNTech) mRNA vaccine has commenced in December 2020.

Pregnant women and their infants, particularly neonates, are at a higher risk for severe SARS-CoV-2 infection. Pregnant women who contract COVID-19 are more likely to be admitted to intensive care unit and require invasive ventilation due to COVID-19 compared to non-pregnant women [5-8]. Furthermore, maternal SARS-CoV-2 infection has been associated with adverse perinatal outcomes including preterm delivery and stillbirth [5-8]. While children have been shown to be more mildly affected by COVID-19 compared with adults, infants are at significantly higher risk for severe disease course as compared to older children [9, 10].

The two SARS-CoV-2 mRNA vaccines of Pfizer and Moderna were shown to elicit a robust immune response among pregnant women [11], coupled with data supporting their safety throughout gestation [12]. In addition to their suggested role in preventing maternal illness, recent studies have demonstrated that antenatal vaccination may lead to transplacental transfer of maternally-derived anti-SARS-CoV-2 antibodies [11, 13-15]. As children, including young infants, are currently not eligible for SARS-CoV-2 vaccination, offering neonatal seroprotection in the early, vulnerable stages of life through maternal immunization is of paramount importance. This principle is well-established for the prevention of other potentially life-threatening respiratory infections such pertussis and influenza [16-20]. Defining the optimal timing for maternal SARS-CoV-2 immunization is crucial to maximize maternofetal antibody transfer and infant protection. Given the high clinical relevance, we aimed to determine the impact of maternal SARS-CoV-2 immunization timing on the efficacy of transplacental antibody transfer.

## Materials and Methods

### Study Population

A prospective study following women admitted for delivery was performed during February-April 2021 at Hadassah Medical Center, a tertiary-care university affiliated hospital in Jerusalem, Israel with over 10,000 deliveries annually. Women who received the SARS-CoV-2 BNT162b2 mRNA vaccine during pregnancy were eligible for this study. The final study cohort included those who received the first vaccine dose at 27-36 weeks gestation. Parturients who delivered prematurely (<37 weeks gestation), multifetal gestations, and those who did not complete the two-dose vaccine series prior to delivery, were excluded. Demographic and clinical data, were collected at the time of enrollment. The institutional review board of the Hadassah Medical Center approved this study (HMO-0064-21).

### Laboratory Methods

Maternal and cord blood sera were collected following delivery after obtaining written informed consent. Spike protein (S) (Liaison SARS-CoV-2 S1/S2 IgG, DiaSorin, Saluggia, Italy) and receptor binding domain (RBD)-specific (Architect SARS-CoV-2 IgG II Quant assay, Abbott Diagnostics, Chicago, USA), IgG levels were evaluated in maternal and cord blood sera. Maternal and cord blood sera were also tested for SARS-CoV-2 RBD IgM (Liaison, DiaSorin, Saluggia, Italy).

For a subset of mother/newborn dyads, neutralizing antibody titers against SARS-CoV-2 were defined using a wild-type SARS-CoV-2 virus microneutralization assay as previously described [21], with minor modifications. Briefly, serial two□fold dilutions of heat inactivated serum samples (starting from 1:10; diluted in DMEM in a total volume of 50 μl) were incubated with an equal volume of viral solution, containing 100 tissue culture infectious dose (TCID50) of SARS-CoV-2 isolate USA-WA1/2020 (NR-52281; obtained from BEI resources), for 1 □hour in a 96-well plate (at 37°C in humidified atmosphere with 5% CO_2_). The serum-virus mixtures (100□µL; 8 replicates of each serum dilution) were then added to a 96-well plate containing a semi□confluent VERO E6 cell monolayer (ATCC CRL-1586; maintained as described [22]). Following 3 days of incubation (at 37°C in a humidified atmosphere with 5% CO_2_), the cells in each well were scored for viral cytopathic effect (CPE). The neutralization titer (NT_50_) was defined as the reciprocal of the highest serum dilution that protected 50% of culture wells from CPE. Positive and negative serum controls, cell control, and a viral back-titration control were included in each assay.

### Statistical analysis

Patient characteristics are described as proportions for categorical variables and medians and interquartile range (IQR) for continuous variables. Antibody levels, neutralizing activity and placental transfer ratios are expressed as medians and IQR. Significance between those vaccinated in early (27-31 weeks) versus late third trimester (32-36 weeks) was assessed using the chi-square test and Fisher ‘s exact test for categorical variables, while the Mann-Whitney U test was used for continuous variables. This comparison was chosen based on prior studies showing that transplacental antibody transfer is most efficient during the third trimester of pregnancy [16]. Correlations were reported using the Pearson ‘s test with the correspondent R and P values. In addition, a multivariable linear regression analysis was performed to assess factors independently associated with neonatal antibody levels. The data were analyzed using Software Package for Statistics and Simulation (IBM SPSS version 24, IBM Corp, Armonk, NY).

### Results

During the study period, samples were collected from 207 parturients who received the SARS-CoV-2 BNT162b2 mRNA throughout gestation and agreed to participate. Of them, 36 (17.4%) were excluded (first vaccine dose <27 weeks [n=26], first vaccine dose >36 weeks [n=3], preterm delivery [n=5], did not complete the two-dose vaccine series prior to delivery [n=1], twin gestation [n=1]). Thus, the final study cohort comprised 171 women including 83 (48.5%) who received the first vaccine dose at early 3^rd^ trimester (27-31 weeks gestation), and 88 (51.5%) who received the first vaccine dose at late 3^rd^ trimester (32-36 weeks gestation) (Figure 1).

**Figure 1:**
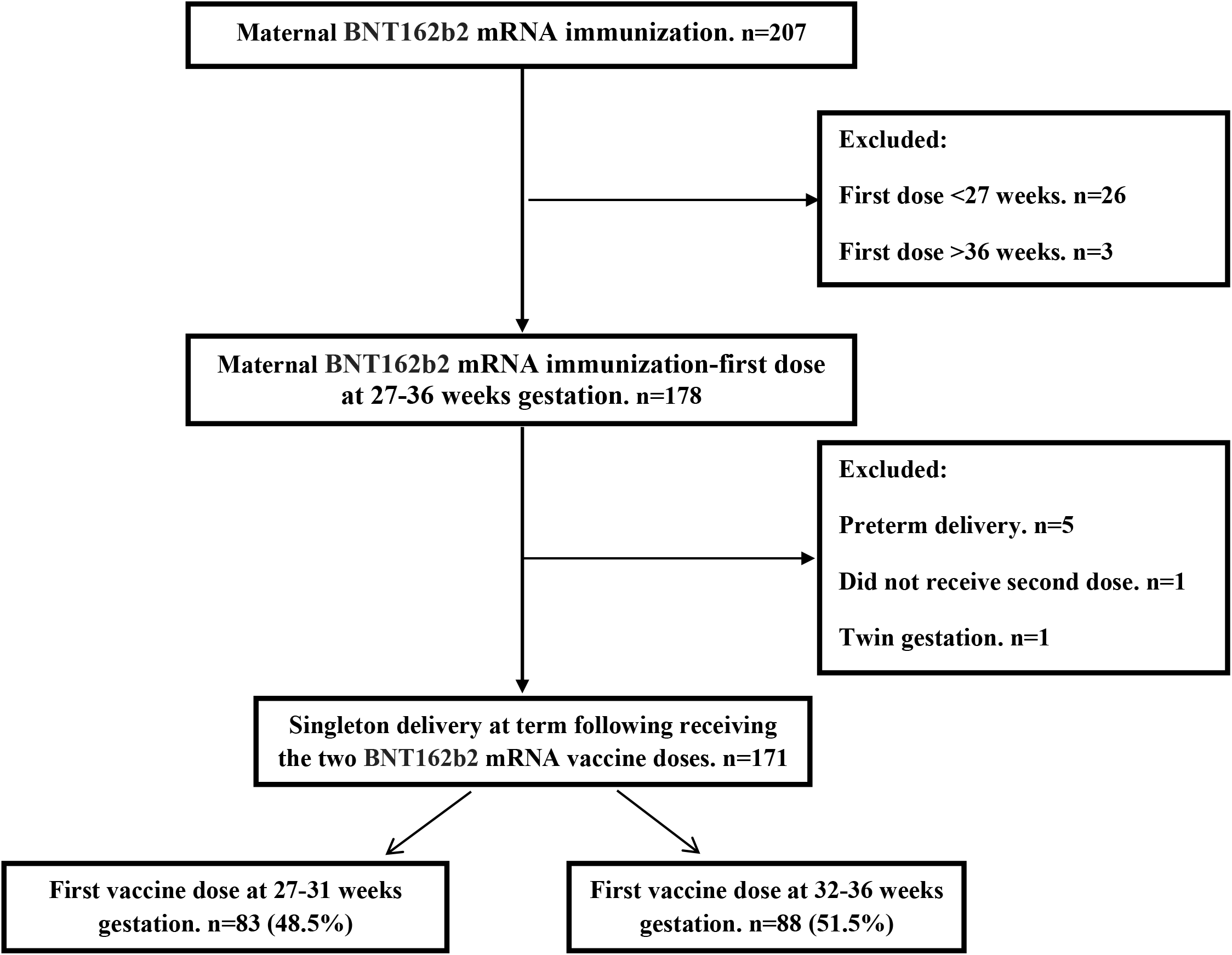
Schematic flow chart of patient inclusion in the study

Maternal and neonatal characteristics are shown in Table S1. Median maternal age was 31 [IQR 27-35] years with a median gestational age of 39^5/7^ [IQR 38^5/7^-40^4/7^] weeks at the time of delivery. Demographic, pregnancy and delivery characteristics did not differ in relation to the time of vaccination. The median time lapsed from the first vaccine dose administration until delivery was 71 [IQR 63-79] and 41 [IQR 34-50] days, in those immunized in the early 3^rd^ trimester and late 3^rd^ trimester, respectively (P<0.001).

All 171 mother-infant pairs were positive for SARS-CoV-2 anti S- and anti-RBD-specific IgG, with a positive correlation between maternal and neonatal cord blood concentrations (r=□0.78; P□<0.001 and r=□0.66; P□<0.001, respectively; Figure 2A, B).

**Figure 2.**
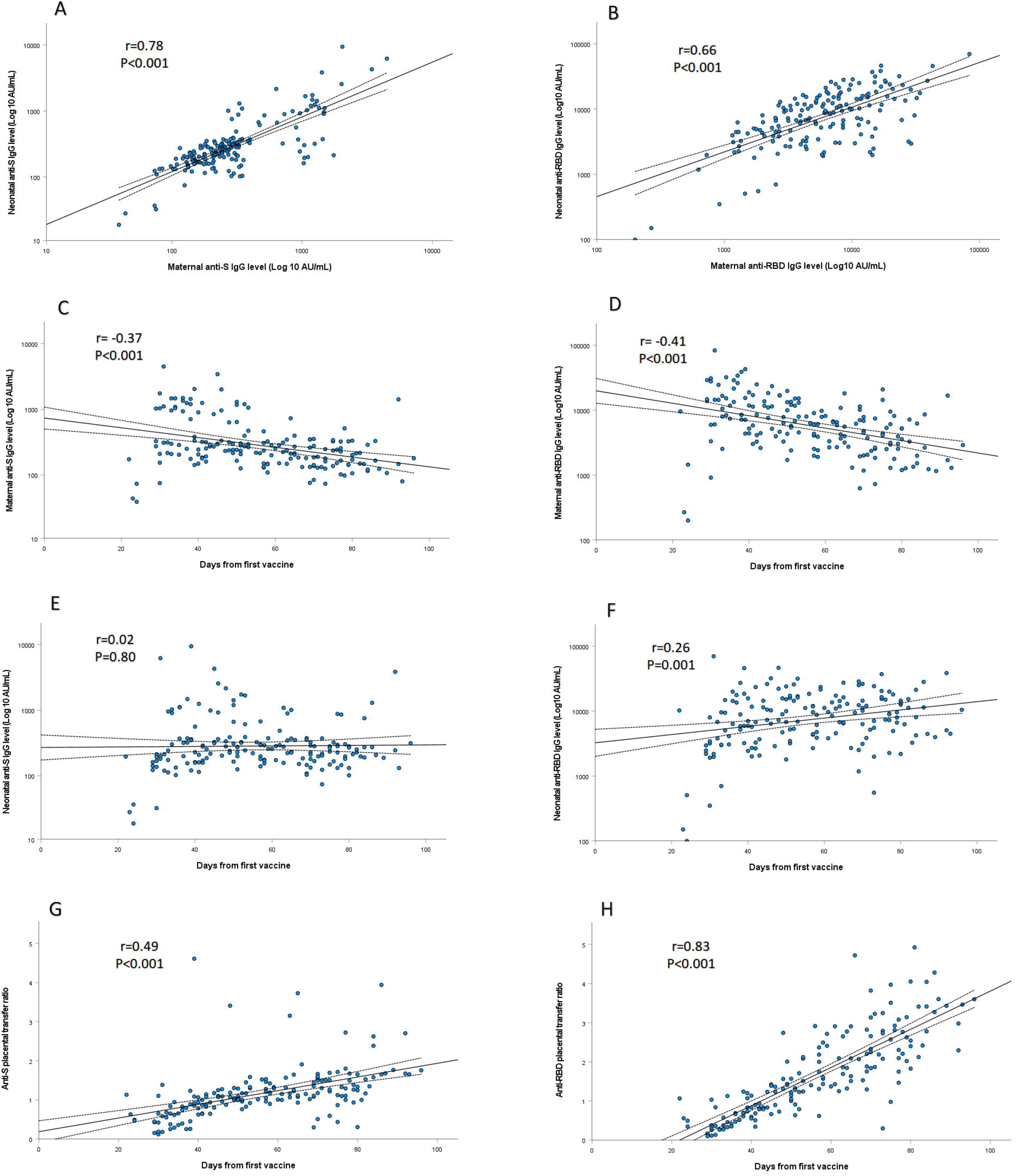
SARS-CoV-2 anti-S (A) and anti-RBD-specific (B) IgG levels in maternal sera were positively correlated to their respective concentrations in cord blood (r=□0.78; P□<0.001 and r=□0.66; P□<0.001, respectively). Maternal anti-S (C) and anti-RBD-specific (D) IgG concentrations were negatively associated with the time lapsed since immunization (r=□-0.37; P□<0.001 and r=□-0.41; P□<0.001, respectively). Neonatal concentrations of anti-S (E) IgG antibodies did not differ in relation to maternal third trimester immunization timing (P=0.80). Anti-RBD-specific (F) IgG concentrations in neonatal sera were positively correlated with increasing time since vaccination (r=□0.26; P=0.001). Placental transfer ratios of anti-S (G) and anti-RBD-specific (H) IgG were directly associated with longer duration since immunization (r=□0.49; P□<0.001 and r=□0.83; P□<0.001, respectively). Correlations, as well as correspondent R and P values were calculated by Pearson ‘s test, as shown in each panel. The dotted lines are the 95% confidence intervals.

SARS-CoV-2 IgM antibodies were not detected in any of the neonates, and were detected in 30 (17.5%) parturients, all of whom were vaccinated late in the 3^rd^ trimester.

Median anti-S and anti-RBD-specific IgG concentrations in maternal sera at the time of delivery were lower in those vaccinated in early 3^rd^ trimester as compared to late 3^rd^ trimester (anti-S IgG: 200 vs. 292 AU/mL, anti-RBD-specific IgG: 3980 vs. 8506 AU/mL, P<0.001 for both comparisons) (Figure 3A, B). There was a negative association between maternal anti-S and anti-RBD-specific IgG concentrations and the time lapsed since immunization (r=□-0.37; P□<0.001 and r=□-0.41; P□<0.001, respectively; Figure 2C, D)

**Figure 3.**
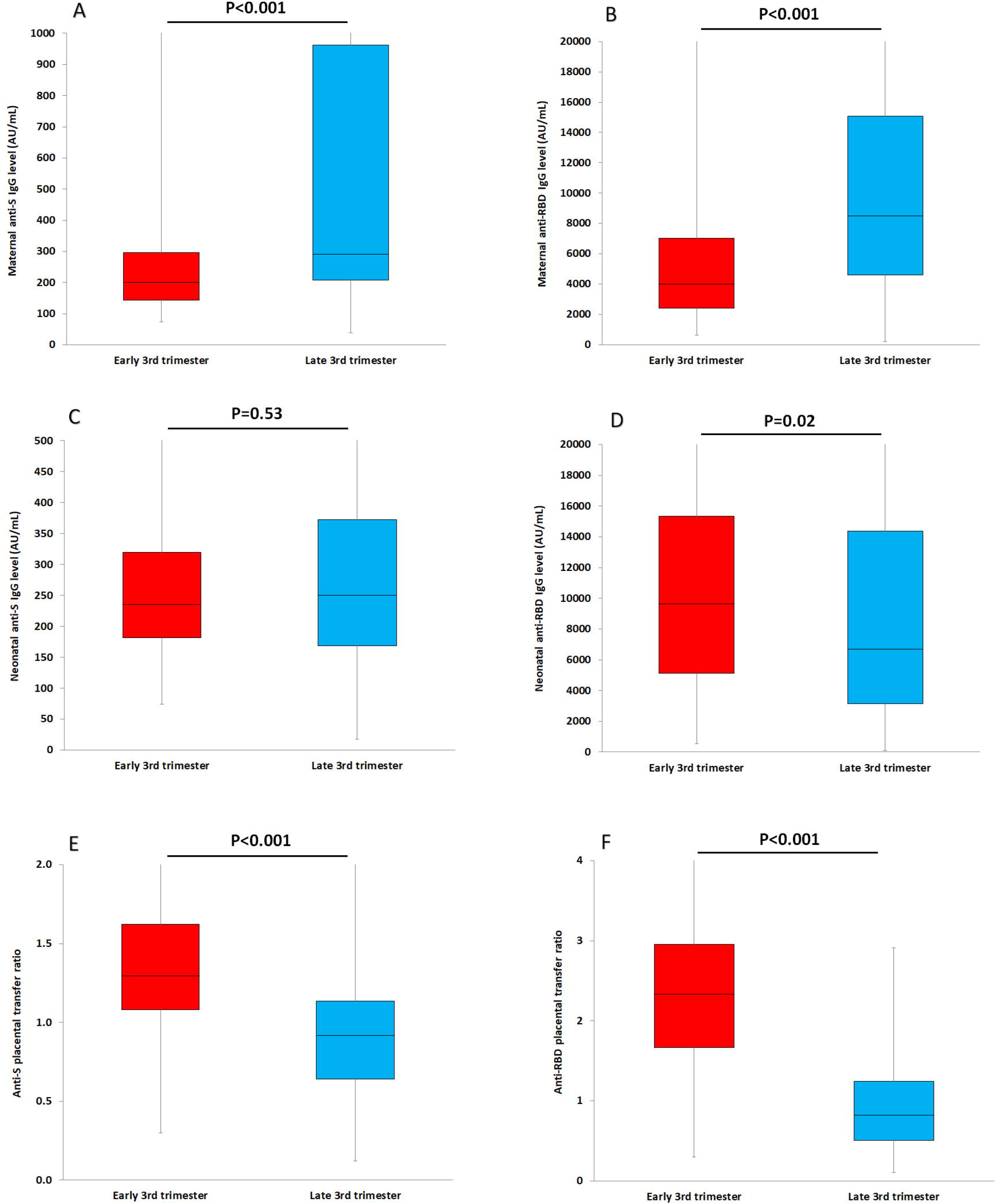
Maternal SARS-CoV-2 anti-S (A) and anti-RBD-specific (B) IgG levels, neonatal anti-S (C) and anti-RBD-specific (D) IgG levels, placental transfer ratios of anti-S (E) and anti-RBD-specific (F) IgG in relation to maternal third trimester immunization timing. Horizontal line represents the median.

Neonatal concentrations of anti-S IgG antibodies did not differ in relation to maternal third trimester immunization timing (Figure 2E, 3C). Anti-RBD-specific IgG concentrations in neonatal sera were significantly higher in those born to mothers vaccinated early in the 3^rd^ trimester as compared to late 3^rd^ trimester (median: 9620 vs. 6697 AU/mL, P=0.02) (Figure 3D), and were positively correlated with increasing time since vaccination (r=□0.26; P=0.001; Figure 2F). The median placental transfer ratios of anti-S and anti-RBD-specific IgG were significantly increased following early 3^rd^ trimester immunization as compared to late 3^rd^ trimester immunization (anti-S ratio: 1.3 vs. 0.9, anti-RBD-specific ratio: 2.3 vs. 0.7, P<0.001 for both comparisons) (Figure 3E, F), and directly associated with longer duration since immunization (r=□0.49; P□<0.001 and r=□0.83; P□<0.001, respectively; Figure 2G, H). Higher proportion of maternal-infant dyads had anti-S and anti-RBD specific IgG placental transfer ratios above 1 following early 3^rd^ trimester immunization as compared to late 3^rd^ trimester immunization (anti-S ratio >1: 83.1% vs. 38.6%, anti-RBD-specific ratio >1: 96.4% vs 38.6%, P<0.001 for both comparison). The sequential decrease in anti-RBD-specific IgG neonatal concentrations and placental transfer ratio in relation to later gestational age at the time of vaccination is shown in Figure 4A, B. Multivariate analysis showed that the only predictor of anti-RBD-specific IgG neonatal concentrations and placental transfer ratio was the duration of time since first vaccine dose administration (β=0.64; P<0.001 and β=0.83; P<0.001, respectively), with no association found with all other maternal, pregnancy and delivery characteristics (see Table S1 for the analyzed characteristic).

**Figure 4.**
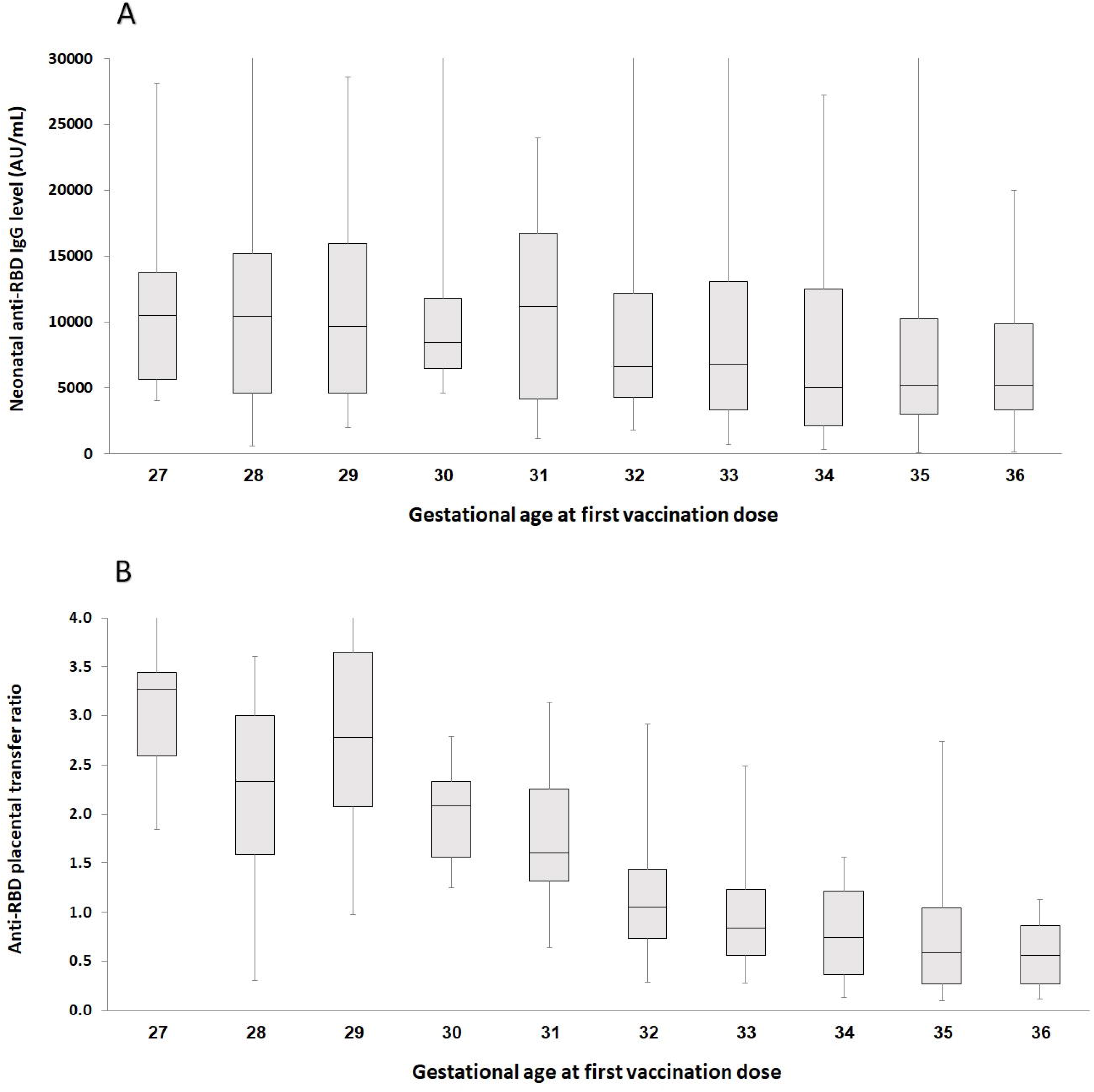
Anti-RBD-specific (A) IgG neonatal concentrations and placental transfer ratio (B) in relation to gestational age at the time of vaccination

Neutralizing SARS-CoV-2 antibodies were assessed in 33 maternal-infant pairs following early 3^rd^ trimester vaccination (n=14) and late 3^rd^ trimester vaccination (n=19). Neonatal sera neutralizing activity (as reflected by NT_50_ values) was positively correlated to maternal sera neutralizing activity (r=□0.43; P=0.01; Figure 5A) and neonatal anti-RBD concentrations (r=□0.68; P<0.001; Figure 5B). Placental transfer ratio of neutralizing SARS-CoV-2 antibodies was significantly higher following early 3^rd^ trimester immunization as compared to late 3^rd^ trimester immunization (median 1.9 vs. 0.8, P<0.001) (Figure 5C), and was positively associated with increasing time from vaccination (r=□0.77; P<0.001; Figure 5D) and anti-RBD placental transfer ratio (r=□0.82; P<0.001; Figure 5E). Neutralizing antibodies placental transfer ratio was above 1 in all 14 (100.0%) mother-neonates dyads vaccinated early in the 3^rd^ trimester and in 7 (36.8%) of those vaccinated in late 3^rd^ trimester (P<0.001).

**Figure 5.**
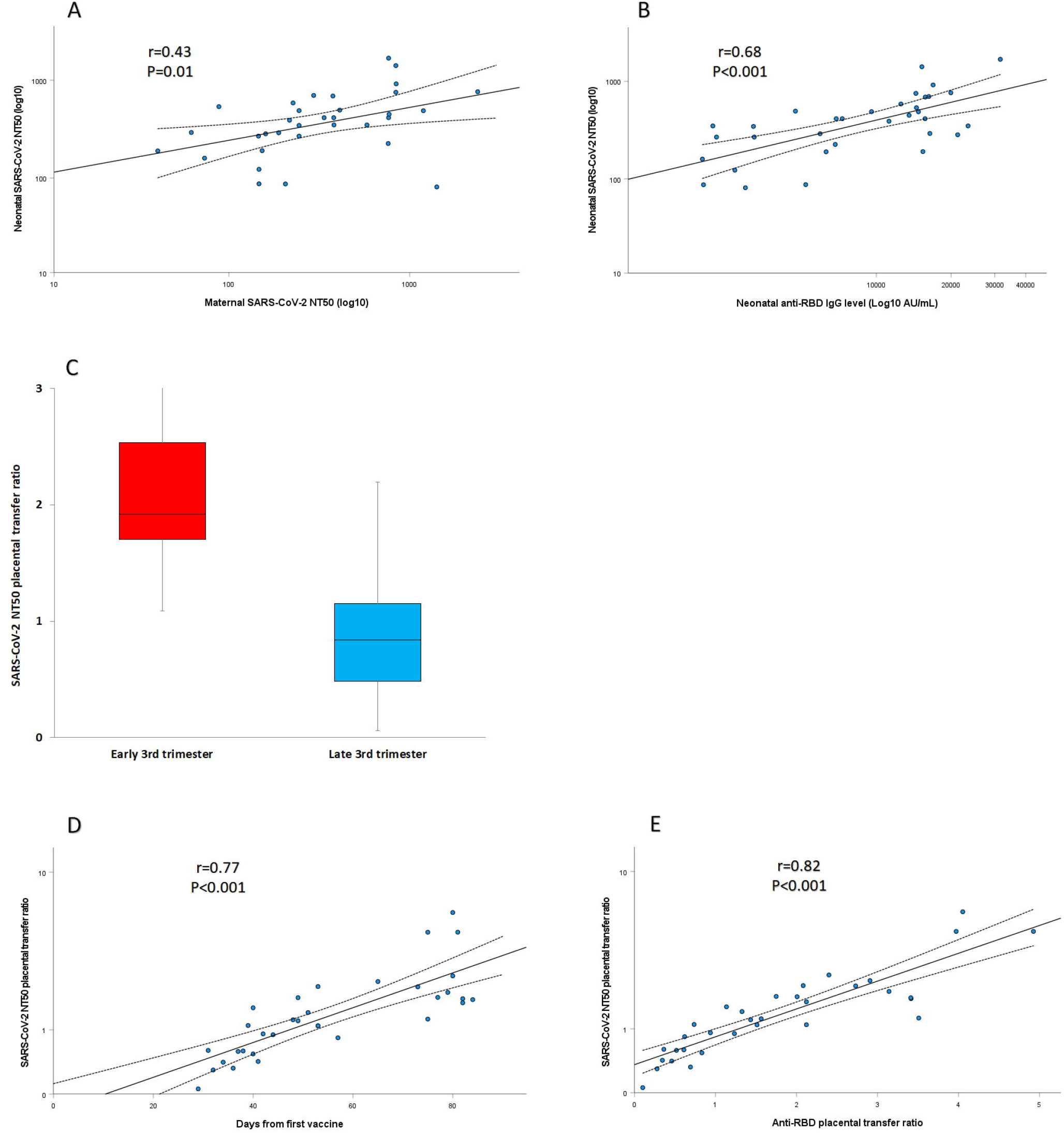
Maternal sera neutralizing activity (A) and neonatal anti-RBD concentrations (B) were positively correlated to neonatal sera neutralizing activity (r=□0.43; P=0.01 and r=□0.68; P<0.001, respectively). Placental transfer ratio of neutralizing following early versus late 3^rd^ trimester immunization (C), horizontal line represents the median. Duration of time since vaccination (D) and anti-RBD placental transfer ratio (E) were positively associated with neutralizing antibodies placental transfer ratio (r=□0.77; P<0.001 and r=□0.82; P<0.001, respectively). Neutralizing activity is reflected by NT_50_ values (see Methods section).

## Discussion

This study investigated the transplacental antibody transfer in 171 mother/newborn dyads, following antenatal SARS-CoV-2 BNT162b2 mRNA vaccination. Early-compared to late third trimester maternal immunization was associated with higher neonatal anti-SARS-CoV-2 antibody levels and serum neutralizing activity. The current study findings may support the role of vaccination early at the third trimester to optimize maternofetal antibody transfer and neonatal seroprotection.

Pregnant women were excluded from the initial trials evaluating the different SARS-CoV-2 vaccines [2, 3]. Nevertheless, while universal recommendations have not yet emerged, given the risk for severe disease course and the accumulating reassuring data for their utilization in the setting of pregnancy [11, 12], the world Health Organization (WHO), the Center for Disease Control and Prevention (CDC), and other agencies advocate offering pregnant women to receive the SARS-CoV-2 vaccine following shared decision making [23-26]. The far-reaching potential to confer neonatal protection against COVID-19 via maternal immunization raises critical questions concerning the optimal timing of antenatal vaccination. Physiologically, transplacental antibody transfer is a dynamic process starting in the second trimester, however, it is most predominant in the third trimester [27, 28]. While for maternal pertussis and influenza vaccination this issue has been extensively studied [16-20], as most women have been previously exposed or immunized against these pathogens, the findings may not apply for the different SARS-CoV-2 vaccines, which prime a de-novo immune response.

In this study, we evaluated pregnant women immunized with the SARS-CoV-2 BNT162b2 mRNA vaccine, with the first dose given between 27-36 weeks gestation. We demonstrated that maternal immunization during the early third trimester (27-31 weeks) yielded higher neonatal antibody concentrations, as compared to late third trimester (32-36 weeks). While the specific antibody titers required for protection have not been fully defined, this enhanced neonatal seroprotection was also evident by the increased neonatal serum anti-SARS-CoV-2 neutralizing antibody levels in those vaccinated earlier. Furthermore, the increment noted in neonatal antibody concentrations and placental transfer ratio in association with earlier gestational age at the time of vaccination, was higher for anti-RBD-specific IgG as compared to anti-S IgG, implying the preferential transfer of the former, correlating with neutralization potency. These observations indicate that neonatal serum is enriched by the placenta with highly functional antibodies. Our results are in accordance with previous reports evaluating the effect of maternal pertussis and influenza immunization, showing augmented transplacental transfer and neonatal antibody levels along with improved clinical outcomes, following early third trimester vaccination [16-20]. While encouraging, future studies should evaluate the kinetics and durability of these passively acquired antibodies in the offspring and their protective effect against with clinical COVID-19-related outcomes.

Aside from the strong correlation between the time lapsed from vaccination and transplacental antibody transfer, we also evaluated the influence of other demographic and clinical characteristics in this regard. While in the setting of third-trimester maternal SARS-CoV-2 infection relatively low placental transfer ratios were previously reported, possibly relating to an altered antibody glycosylation profile [29], we observed higher and satisfactory maternofetal antibody transfer. In addition, although mothers with COVID-19 carrying a male fetus were shown by Bordt et al. to exhibit a diminished immune response and compromised neonatal seroprotection [30], this was not observed in our cohort following maternal vaccination. Furthermore, the lack of detrimental effect of maternal anthropometric parameters and the prenatal administration of anti-D immunoglobulin, on the level of anti-SARS-CoV-2 antibodies conveyed to the offspring are other reassuring findings.

The major strength of our study is its relatively large cohort size, which allowed for a robust analysis of the superiority of early third trimester immunization. Nevertheless, this study has several caveats. First, the observational single-center design of the study may limit the generalizability of our findings. Second, the effects of maternal immunization in early pregnancy as well as of the impact of other SARS-CoV-2 vaccines on transplacental antibody transfer dynamics remain to be determined. The former could provide maternal protection against COVID-19 through the longer course of pregnancy and may benefit preterm infants. Finally, vaccine-induced maternally-derived antibodies might blunt the infant humoral immune response to future vaccination; while the clinical significance of this interference effect is largely unknown [28], it should be acknowledged and further explored.

## Conclusions

The current study results indicate that early third trimester immunization has the potential to maximize maternofetal transplacental antibody transfer thereby affording adequate seroprotection during early infancy. As the strategy of maternal SARS-CoV-2 immunization is gaining support, there is a critical need for further scientific evidence to inform the ideal timing for vaccination during pregnancy that would provide mothers and neonates with the highest clinical protection against COVID-19. As the extent of SARS-CoV-2 community spread declines in several parts of the globe, optimizing neonatal immunity may be more heavily weighted by immunization policy makers.

## Data Availability

Individual-level data will not be made publicly available with this Article. Requests for sharing of deidentified individual-level participant data for scientific research can be directed to the corresponding authors. All proposals will be subject to scientific review and institutional review board approval at Hadassah Medical Center.

## Authors ‘ contributions

Dr Rottenstreich and Prof. Wolf had full access to all of the data in the study and takes responsibility for the integrity of the data and the accuracy of the data analysis.

Concept and design: Porat, Rottenstreich, Wolf.

Acquisition, analysis, or interpretation of data: All authors.

Drafting of the manuscript: Rottenstreich, Zarbiv, Porat, Kleinstern, Oiknine-Djian, Wolf.

Laboratory analyses: Oiknine-Djian, Vorontsov, Wolf.

Statistical analysis: Rottenstreich, Kleinstern.

All authors read and approved the final manuscript.

## Acknowledgments

We thank Dr. Doron Kabiri, Dr. Shlomi Yahalomi, Prof. Yosef Ezra and Dr. Roy Alter for their assistance in patients ‘ enrollment. We also thank Rimma Barsuk and Yulia Yachnin for their technical assistance.

## Data sharing

Individual-level data will not be made publicly available with this Article. Requests for sharing of deidentified individual-level participant data for scientific research can be directed to the corresponding author. All proposals will be subject to scientific review and institutional review board approval at Hadassah Medical Center.

## Legends for tables and figures

**Table S1**

Maternal and neonatal characteristics among SARS-CoV-2 BNT162b2 immunized pregnant women in relation to vaccination timing

