## Supplementary material for "Early versus late third trimester maternal SARS-CoV-2 BNT162b2 mRNA immunization maximizes transplacental antibody transfer and neonatal neutralizing antibody levels": Table S1

Maternal and neonatal characteristics among SARS-CoV-2 BNT162b2 immunized pregnant women in relation to vaccination timing

| **Late 3^rd^ trimester vaccination**  **n=88 (51.5%)** | **Early 3^rd^ trimester vaccination**  **n=83 (48.5%)** | **Characteristics** |
| --- | --- | --- |
| 32 [29-35] (32)  17 (19.3%) | 30 [27-35] (31)  20 (24.1%) | Age (years)  >35 years |
| 3 [2-5] (3)  24 (27.3%) | 2 [1-4] (3)  28 (33.7%) | Parity  Nulliparous |
| 74 [66-82] (76) | 75 [69-84] (76) | Maternal weight (kg) |
| 28 [26-31] (28) | 28 [26-31] (29) | Maternal body mass index (kg/m^2^) |
| 39^6/7^ [38^5/7^-40^6/7^] (39^6/7^) | 39^5/7^ [38^6/7^ -40^3/7^] (39^4/7^) | Gestational age at delivery (weeks) |
| 7 (8.0%) | 6 (7.2%) | Gestation diabetes mellitus |
| 2 (2.3%) | 1 (1.2%) | Gestational hypertensive disorders |
| 6 (6.8%) | 9 (10.8%) | Received antenatal anti D immunoglobulin |
| -  -  -  -  -  27 (30.7%)  26 (29.5%)  15 (17.0%)  9 (10.2%)  11 (12.5%) | 15 (18.1%)  18 (21.7%)  18 (21.7%)  17 (20.5%)  15 (18.1%)  -  -  -  -  - | Gestational age at 1^st^ dose immunization (weeks)  27  28  29  30  31  32  33  34  35  36 |
| 41 [34-50] (42) | 71 [63-79] (71) | 1^st^ vaccine dose-to-delivery interval (days) |
| 20 [13-29] (21) | 50 [42-58] (50) | 2^nd^ vaccine dose-to-delivery interval (days) |
| 81 (92.0%)  5 (5.7%)  2 (2.3%) | 77 (92.8%)  3 (3.6%)  3 (3.6%) | Mode of delivery  Vaginal  Pre-labor cesarean  In-labor cesarean |
| 3440 [3051-3700] (3374) | 3275 [2985-3730] (3345) | Neonatal Birthweight (grams) |
| 50 (56.8%) | 39 (47.0%) | Male gender (%) |

All continuous variables are expressed as medians [interquartile range] (means).
